# Discrete mechanistic pathways underlying genetic predisposition to atrial fibrillation are associated with different intermediate cardiac phenotypes and risk of cardioembolic stroke

**DOI:** 10.1101/2024.07.10.24310244

**Authors:** PR Gajendragadkar, A Von Ende, F Murgia, A Offer, CF Camm, RS Wijesurendra, B Casadei, JC Hopewell

**Affiliations:** Clinical Trial Service Unit and Epidemiological Studies Unit, Nuffield Department of Population Health, University of Oxford; Division of Cardiovascular Medicine, Radcliffe Department of Medicine, University of Oxford

**Keywords:** Atrial fibrillation, ischaemic stroke, genetics, big data

## Abstract

**Background:** Genome-wide association studies (GWAS) have clustered candidate genes associated with atrial fibrillation (AF) into biological pathways reflecting different pathophysiological mechanisms. We investigated whether these pathways associate with distinct intermediate phenotypes and confer differing risks of cardioembolic stroke.

**Methods:** Three distinct subsets of AF-associated genetic variants, each representing a different mechanistic pathway, i.e., the cardiac muscle function and integrity pathway (15 variants), the cardiac developmental pathway (25 variants), and the cardiac ion channels pathway (12 variants), were identified from a previous AF GWAS. Using genetic epidemiological methods and large-scale datasets such as UK Biobank, deCODE, and GIGASTROKE, we investigated the associations of these pathways with AF-related cardiac intermediate phenotypes, which included ECG parameters (∼16,500 ECGs), left atrial and ventricular size and function (∼36,000 cardiac MRI scans), and relevant plasma biomarkers (NT-proBNP, ∼70,000 samples; high-sensitivity troponin I and T, ∼87,000 samples), as well as with subtypes of ischaemic stroke (∼11,000 cases).

**Results:** Genetic variants representing distinct AF-related mechanistic pathways had significantly different effects on several AF-related phenotypes. In particular, the muscle pathway was associated with a longer PR interval (*P* for heterogeneity between pathways [*P*_het_] = 1 × 10^-10^), lower LA emptying fraction (*P*_het_ = 5 × 10^-5^), and higher NT-proBNP (*P*_het_ = 2 × 10^-3^) per log-odds higher risk of AF compared to the developmental and ion channel pathways. By contrast, the ion channel pathway was associated with a lower risk of cardioembolic stroke (*P*_het_ = 0.04 in European, and 7 × 10^-3^ in multi-ancestry populations) compared to the other pathways.

**Conclusions:** Genetic variants representing specific mechanistic pathways for AF are associated with distinct intermediate cardiac phenotypes and a different risk of cardioembolic stroke. These findings provide a better understanding of the aetiological heterogeneity underlying the development of AF and its downstream impact on disease and may offer a route to more targeted treatment strategies.

## INTRODUCTION

Atrial fibrillation (AF) is the electrical end-point of radically different pathophysiological pathways^1, 2^. Highlighting this complexity, linkage and family studies have associated AF with rare variants in genes encoding not just ion channels^3, 4^, but transcription factors^5^, hormones^6^, and more recently, cardiac structural proteins^7–11^. In line with these findings, pathway and functional enrichment analyses from a large genome-wide association study (GWAS) have suggested that many of the genes annotated to AF-associated loci can be grouped into mechanistic pathways reflecting cardiac and skeletal muscle function and integrity, developmental events, hormone signalling, angiogenesis and myocardial ion channels^12^.

In individuals with AF, a number of biomarkers and intermediate phenotypes have shown associations with subsequent AF-related outcomes, such as stroke^13–15^. These findings may provide opportunities for improving the management and risk stratification of patients with AF, as well as for repurposing drugs that are already available for treating other diseases^12^. However, there remains uncertainty in our understanding of whether AF-associated mechanistic pathways are associated with distinct cardiac phenotypes and differing risks of severe complications.

Using clusters of genetic variants representing different AF-associated mechanistic pathways^12^, we investigated associations between genetically-predicted AF pathways and AF-related intermediate phenotypes, encompassing cardiac electrical, structural, and functional characteristics (ECG parameters^16–18^, left atrial [LA] size and function^19–21^, left ventricular [LV] function^22, 23^, N-terminal pro B-type natriuretic peptide [NT-proBNP]^24, 25^, and high-sensitivity cardiac troponins^25–27^), and examined their relevance to risk of stroke.

## METHODS

A summary of the study design is shown in **Supplementary Figure 1**.

### Genetically-predicted mechanistic pathways associated with risk of AF

A total of 142 independent genome-wide significant variants (*P* < 5 × 10^-8^) were previously identified in a large AF GWAS meta-analysis, referred to as the comprehensive 142-variant set hereafter, which captures broader genetic predisposition to AF^12^. In the same study, these variants were annotated to functional genes using a stringent multi-step process^12^ in the following order of priority: whether they were protein-altering, or in high linkage disequilibrium with protein-altering variants; had expression levels that were associated and co-localised with AF-associated loci across all tissue using Genotype-Tissue Expression (GTEx) consortium data; were identified by DEPICT (Data-driven Expression Prioritized Integration for Complex Traits); or were the nearest gene to index variant in a locus. A further step grouped functional candidate genes (whenever possible) into mechanistic pathways in which they were likely to be involved (e.g. muscle, developmental, ion channel)^12^. Further details on the locus to gene and pathway annotation processes used in the GWAS^12^ are found in the **Supplementary Methods**.

In the present study, we used independent genetic variants that had been annotated to three distinct AF-related mechanistic pathways. The ‘muscle pathway’ comprised variants annotated to genes involved in cardiac and skeletal muscle function and integrity (15 genetic variants, annotated to 14 distinct genes e.g. *MYH6*, *PKP2*, *TTN*). The ‘developmental pathway’ comprised variants annotated to genes involved in cardiac developmental events (25 genetic variants, annotated to 12 distinct genes e.g. *GATA4*, *NKX2-5*, *PITX2*, *TBX5*). The ‘ion channel pathway’ comprised variants annotated to genes involved in cardiac ion channels (12 genetic variants, annotated to 8 distinct genes e.g. *HCN4*, *KCNH2*, *KCNJ5*, *SCN5A*). Of the comprehensive 142-variant set, 52 variants were annotated to the genes likely to be involved in one of these three discretely defined non-overlapping pathways.

The AF GWAS meta-analysis^12^ also identified variants annotated to genes likely to be involved in calcium handling (4 variants), angiogenesis (1 variant) and hormone signalling (4 variants). These potential pathways were not included in the present study as they were relatively non-specific and comprised multiple heterogeneous biological pathways. For example, the hormone signalling pathway identified genes associated with oestrogen receptors, thyroid receptors and insulin-like growth factor receptors. Further details are provided in **Supplementary Tables 1 - 4**.

### Study populations and outcome definitions

Details of the datasets used in this study are listed in **Supplementary Table 5**.

#### Atrial fibrillation

Genetic associations with AF were generated directly in UK Biobank^28, 29^ in 339,189 unrelated white British participants whose genetic samples passed bioinformatic quality control using logistic regression with adjustments for sex, genotype array, and 7 principal components of population structure. We identified 30,631 cases of AF or flutter, defined using electronic healthcare records with details of definitions in **Supplementary Table 6**. Further details of phenotype codes utilised to define co-morbidities are listed in **Supplementary Table 7**.

#### ECG parameters

As with AF, genetic associations with ECG parameters were generated directly in UK Biobank using logistic regression with adjustments for sex, genotype array, and 7 principal components of population structure. Resting 12-lead ECGs and automated measurements (using Cardiosoft v6 program, GE) were available in 16,508 participants, and analyses of these data were conducted after excluding individuals with AF on ECG, or AF identified as described above.

#### Cardiac MRI parameters

Genetic associations with cardiac MRI imaging data including LA structure and function^30^ (35,658 individuals of European ancestry) and LV structure and function^31^ (36,041 individuals of European ancestry), both regardless of history of AF, were based on existing summary data derived from UK Biobank. The UK Biobank cardiac MRI protocol has been reported elsewhere^32^. In brief, LA parameters were derived from automated processing algorithms; volumes were indexed to Du Bois formula calculated body surface area and cardiac volume-time curves created to derive active and passive LA emptying fractions^30^. LV parameters were provided by UK Biobank, corrected for known bias from the automated algorithm and indexed to Mosteller formula calculated body surface area^31^.

#### NT-proBNP

Genetic associations with NT-proBNP were based on summary data including 33,043 individuals from the UK Biobank Pharma Proteomics Project (UKB-PPP)^33^ and 35,559 Icelandic individuals from the deCODE + Icelandic cancer project^34^. These associations were further validated in summary data including 3,301 healthy blood donors from the INTERVAL study^35, 36^ and a GWAS of 3,394 individuals with cardiovascular risk factors from the IMPROVE study^37, 38^. Assays for NT-proBNP included Olink (UKB-PPP, IMPROVE) and SomaScan (deCODE + ICP, INTERVAL).

#### High-sensitivity troponin I and T (hsTnI, hsTnT)

Genetic associations with hsTnI were based on summary data including 48,115 individuals from the Trøndelag Health Study (HUNT) and Generation Scotland Family Health Study (GS:FHS)^39^ and in 14,336 individuals of diverse ancestry (89% European, 11% African) across a number of cohorts (including ARIC) but excluding individuals with baseline coronary heart disease and heart failure as described by Yang *et al*^40^. Genetic associations with hsTnT were based on summary data including 24,617 individuals (76% European, 15% African, 7% Hispanic, 3% Asian) across a number of cohorts (including ARIC, CHS, MESA) but excluding individuals with baseline coronary disease and heart failure^40^.

#### Ischaemic stroke

Genetic associations with stroke outcomes were based on summary data from the GIGASTROKE GWAS meta-analysis^41^. Contributing studies defined the aetiology of stroke as per the TOAST (Trial of Org in 10172 Acute Stroke Treatment) classification system^42^. In the present study, we examined associations with cardioembolic stroke (10,804 European ancestry; 12,790 multi-ancestry) as a downstream consequence of AF, and small-vessel stroke as a negative control due to its different risk factor profile (6,811 European; 13,620 multi-ancestry).

### Statistical analysis

#### Primary analyses

Ratio estimates for each individual variant were determined by dividing the gene-outcome association (for each outcome of interest as described above) by the association with AF (taken from the published GWAS for AF^12^ after exclusion of UK Biobank participants to reduce the potential for bias). For each specific pathway, the individual variant ratio estimates were then combined using fixed-effects (assuming a single common effect) inverse variance weighted methods, with resulting estimates corresponding to a unit increase in log-odds risk of AF (approximately a 2.7-fold higher risk).

Heterogeneity between estimates from different pathways was assessed using Cochran’s Q statistic and Wald tests to examine pair-wise differences between individual pathways. Nominal significance was defined as a *P*-value <0.05, with more robust statistical significance defined as a *P*-value <0.01 to account for within-study multiple testing.

#### Sensitivity analyses

We conducted sensitivity analyses to assess the robustness of the results to underlying methodological assumptions (based on a Mendelian randomisation [MR] instrumental variable framework) including: inverse variance weighted random effects (allowing for heterogeneity between instrumental variables); weighted median (in which up to 50% of the information is permitted to be from invalid instrumental variables); weighted mode (which allows the majority of information to be invalid provided the most common MR effect estimate is a consistent estimate of the true MR effect); and MR-Egger (in which all genetic variants are permitted to be invalid instrumental variables, provided that the pleiotropic and risk factor effects of the variants are independently distributed)^43, 44^. MR estimates from each pathway were also estimated after exclusion of outlier variants identified using the MR-PRESSO outlier detection method^45^. ‘Leave-one-out’ analyses were performed to test whether the effects of the pathways on cardioembolic stroke were sensitive to individual variants.

Additional post-hoc analyses were also undertaken to estimate the direct effects of the pathways on outcomes showing heterogeneity between pathways (i.e. cardioembolic stroke), independent of their associations with ECG parameters, LA parameters, and markers of LV dysfunction. To further confirm that the scores were unrelated to conventional risk factors, within UK Biobank, participant-level weighted AF risk scores were calculated for each of the pathways and the characteristics of the top and bottom quintiles of score compared.

All analyses were performed using SAS (v 9.4, SAS Institute) and R (v 4.3.1, The R Foundation for Statistical Computing).

### Standard protocol approvals, registrations, and patient consents

UK Biobank data were used from the UK Biobank Resource under Application Number 14568. All procedures and data collection in UK Biobank were approved by the UK Biobank Research Ethics Committee (reference number 11/NW/0274) with participants providing full written informed consent for participation and subsequent use of their data for approved applications. Publicly available non-identifiable summary statistics were used in this study for which the multiple contributory studies to the various datasets had their own consent procedures.

## RESULTS

### Comparable risks of AF unrelated to demographics or co-morbidities across mechanistic pathways of genetic predisposition to AF

The estimated associations of AF of the three discrete pathways (‘muscle’; 15 genetic variants, ‘developmental’; 25 variants, and ‘ion channel’; 12 variants) are based on the individual genetic variants’ associations in the AF GWAS meta-analysis (excluding UK Biobank)^12^. As a validation step, we estimated the effects of these pathways on AF in the UK Biobank population. As anticipated, the pathways conferred comparable risks of AF (*P* for heterogeneity 0.72; **Supplementary Figure 2**).

To investigate if the different pathways were associated with risk factors for AF, individual weighted genetic AF risk scores for each of the pathways were calculated in UK Biobank. Participants were split into quintiles of AF risk score and characteristics of the top versus bottom 20% compared (**Supplementary Table 8**). As expected, other than risk of AF, no meaningful differences in demographics or co-morbidities were noted.

### Differing associations with cardiac intermediate phenotypes across mechanistic pathways for same genetic predisposition to AF

#### Surface ECG parameters

In 16,508 participants from UK Biobank with no history of AF, the genetically-predicted AF pathways were associated with different effects on P-wave duration, and PR interval (*P* for heterogeneity 0.03, and 1 × 10^-10^ respectively; **Figure 1**). In particular, the developmental and ion channel pathways were associated with a shorter P-wave duration, but the muscle pathway was not (**Figure 1**). Conversely, the muscle pathway was associated with a *longer* PR interval (by 5.58 ms, 95% CI: 2.59 to 8.58, *P* = 3 × 10^-4^ per log-odds higher genetically-predicted risk of AF), whilst the ion channel pathway was associated with a *shorter* PR interval (by 8.64 ms, 95% CI: -11.53 to -5.75, *P* = 4 × 10^-9^ per log-odds higher genetically-predicted risk of AF; **Figure 1**). This pattern remained the same after adjusting the ECG PR interval for the P-wave duration, (*P* for heterogeneity 3 × 10^-10^; **Supplementary Figure 3**).

**Figure 1.**
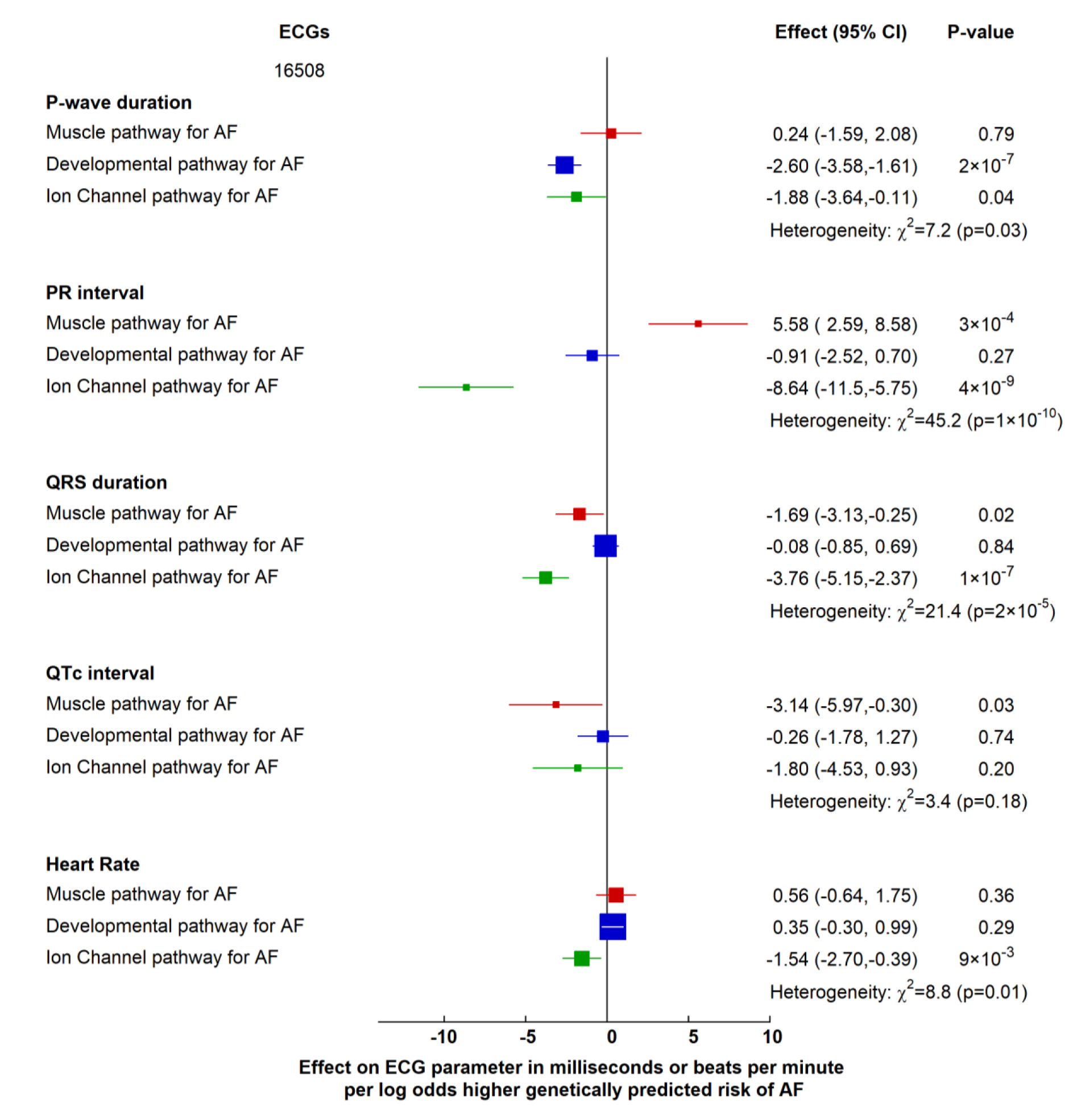
Effects of genetically predicted AF biological pathways on surface ECG parameters in UK Biobank participants. Effect of genetically predicted AF biological pathways on ECG parameters (in milliseconds) or heart rate (in beats per minute) in a UK Biobank cohort of 16,508 participants without a history of AF. Boxes represent point estimates of effect per log-odds higher genetically predicted risk of AF, with their size inversely proportional to variance and solid lines representing 95% confidence intervals (CI). Effect sizes were calculated using inverse variance weighted methods, and heterogeneity using Cochran Q.

There were small but significant differences between the associations with QRS duration for the three pathways (*P* for heterogeneity 2 × 10^-5^; **Figure 1**), but not with corrected QT interval (*P* for heterogeneity 0.18; **Figure 1**). There were differences between the associations of the pathways with heart rate (*P* for heterogeneity 0.01; **Figure 1**), with a log-odds higher genetically-predicted risk of AF via the ion channel pathway being associated with a lower heart rate (-1.54 bpm 95% CI: -2.70 to -0.39, *P* = 9 × 10^-3^), whereas other pathways showed no association.

By contrast, overall AF genetic susceptibility (i.e., as conferred by the 142-variant set) was not associated with differences in PR interval duration (*P* = 0.33; **Supplementary** Figure 4) but was associated with a shorter P-wave (-1.25 ms; 95% CI: -1.79 to -0.71; *P* = 6 × 10^-6^) and QRS duration (-0.95 ms; 95% CI: -1.37 to -0.53, *P* = 1 × 10^-5^) per unit log-odds higher risk of AF.

These findings indicate that mechanistic pathways of genetic predisposition to AF have different associations with ECG parameters (most strikingly for the PR interval), which may not be apparent when overall genetic AF susceptibility is considered.

#### Left atrial size and function

We then investigated the association of the pathways with LA structure and function using GWAS summary data from 35,658 UK Biobank participants who underwent a cardiac MRI scan^30^. Whilst each AF pathway was similarly associated with larger maximum LA volumes (*P* for heterogeneity 0.33; **Figure 2A**), the magnitude of the associations with minimum LA volume differed across the pathways (*P* for heterogeneity 0.02; **Figure 2A**). Specifically, the muscle pathway was associated with a larger minimum LA volume compared with the developmental (*P* = 7 × 10^-3^) and with the ion channel pathway (*P* = 0.04). The 142-variant set for overall AF susceptibility was associated with larger maximal and minimum LA volumes (**Supplementary Figure 5A**).

**Figure 2.**
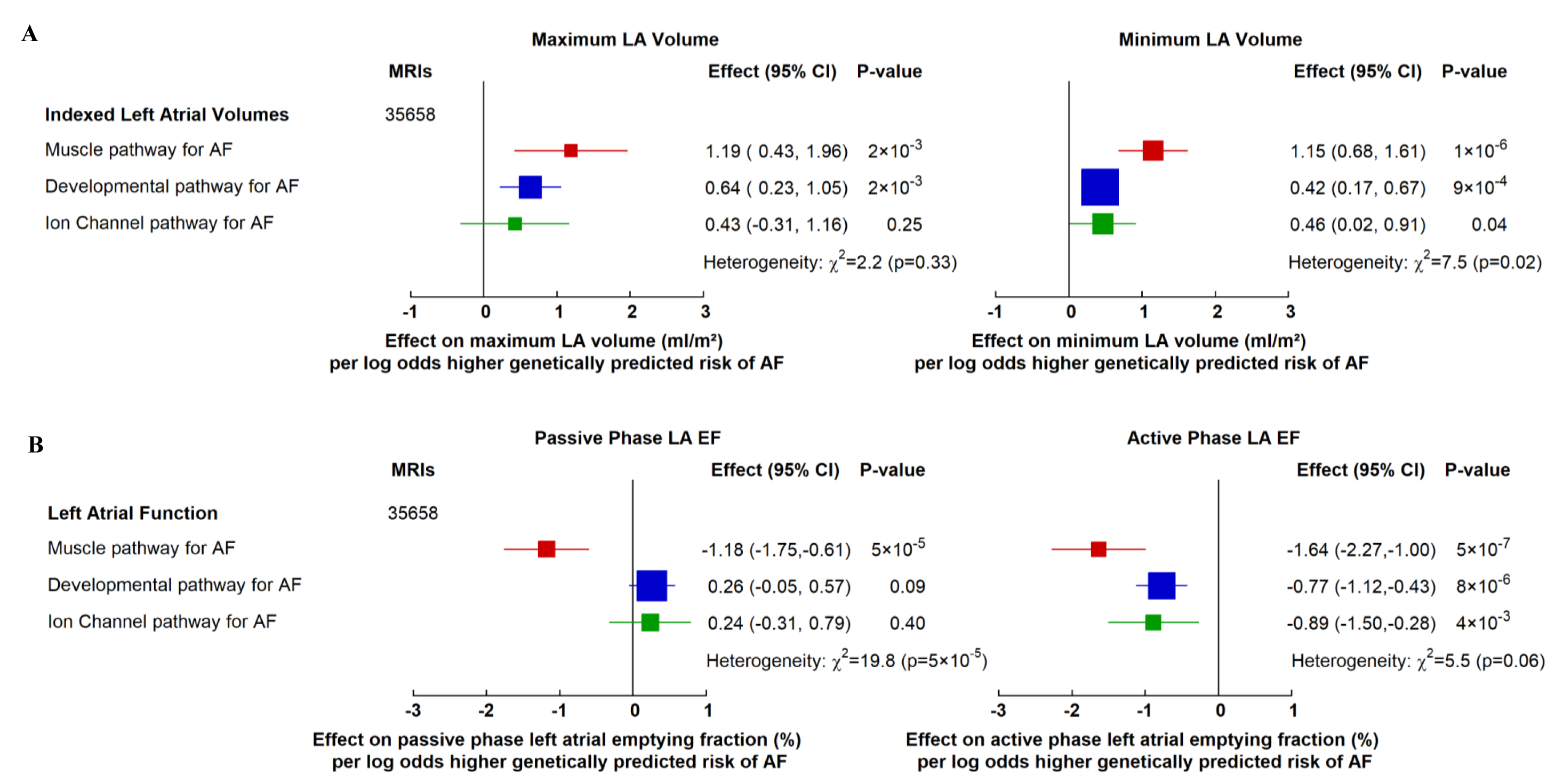
Effects of genetically predicted AF biological pathways on left atrial structure and function in UK Biobank participants. Effect of AF pathways on (**A**) indexed left atrial (LA) volumes in ml/m^2^ and (**B**) LA passive and active phase emptying fractions from 35,658 cardiac MRIs in UK Biobank cohort. Boxes represent point estimates of effect per log odds higher genetically predicted risk of AF, with their size inversely proportional to variance and solid lines representing 95% confidence intervals (CI). Effect sizes were calculated using inverse variance weighted methods from summary GWAS data, and heterogeneity using Cochran Q.

When assessing LA function, we used previously defined passive and active phase LA emptying fraction measurements from cardiac MRI^30^. There were differences in the effects observed between different pathways for passive phase LA emptying fraction (*P* for heterogeneity 5 × 10^-5^). Specifically, a higher genetically-predicted risk of AF *via* the muscle pathway was associated with a lower passive phase LA emptying fraction (-1.18%; 95% CI: -1.75 to -0.61, *P* = 5 × 10^-5^), but no association was observed with the other pathways (**Figure 2B**). The comprehensive 142-variant set for AF susceptibility was not associated with passive phase LA emptying fraction (**Supplementary Figure 5B**).

By contrast, each of the three mechanistic pathways and the 142-variant set were associated with a lower active phase LA emptying fraction (**Figure 2B, Supplementary Figure 5B**), and whilst the effect of the muscle pathway was the most extreme (as observed for the passive phase of LA emptying fraction), there was no significant difference between the pathways (*P* for heterogeneity 0.06; **Figure 2B**).

In summary, different mechanistic pathways of genetic predisposition to AF had different associations with LA structure and function. Amongst these, the muscle pathway was associated with larger LA size and more impaired LA function compared with the other pathways.

#### Left ventricular function

Left ventricular dysfunction and AF share a complex bi-directional relationship. To investigate whether biologically-informed pathways underpinning AF risk were differentially linked to imaging biomarkers of LV function, we examined their associations with LV ejection fraction (LVEF) using GWAS summary data from 36,041 UK Biobank participants who underwent a cardiac MRI scan^31^.

None of the individual pathways were associated with LVEF, and there were no significant differences between pathways (**Supplementary Table 9**). The muscle pathway was associated with a 0.65 ml/m^2^ lower indexed LV stroke volume (95% CI: -1.18 to -0.12; *P* = 0.02), but associations did not differ significantly between the pathways (*P* for heterogeneity 0.14; **Supplementary Table 9**). The 142-variant set was associated with a 0.17% lower LV ejection fraction (95% CI: -0.23 to -0.004; *P* = 0.04) and a 0.20 ml/m^2^ lower indexed LV stroke volume (95% CI: -0.35 to -0.04; *P* = 0.01).

#### NT-proBNP

We also examined associations with NT-proBNP using summary statistics from a cohort of 33,043 individuals from the UKB-PPP^33^ (mean age 59 years and similar to the whole UK Biobank cohort in terms of co-morbidities) and a cohort of 35,559 Icelandic individuals from the deCODE + Icelandic cancer projects^34^ (mean age 55 years with a variety of co-morbidities).

The biological pathways showed different associations with NT-proBNP (*P* for heterogeneity 2 ×10^-3^ and 2 ×10^-5^ for UKB-PPP and deCODE + ICP respectively; **Figure 3**). In these two large cohorts, only the muscle pathway was associated with higher levels of NT-proBNP (0.13 standard deviation (SD) unit higher; 95% CI: 0.06 to 0.20, *P* = 5 × 10^-4^ in UKB-PPP and 0.24 SD unit higher; 95% CI: 0.16 to 0.32, *P* = 4 × 10^-9^ in deCODE + ICP), with no associations noted for the other pathways (**Figure 3**).

**Figure 3.**
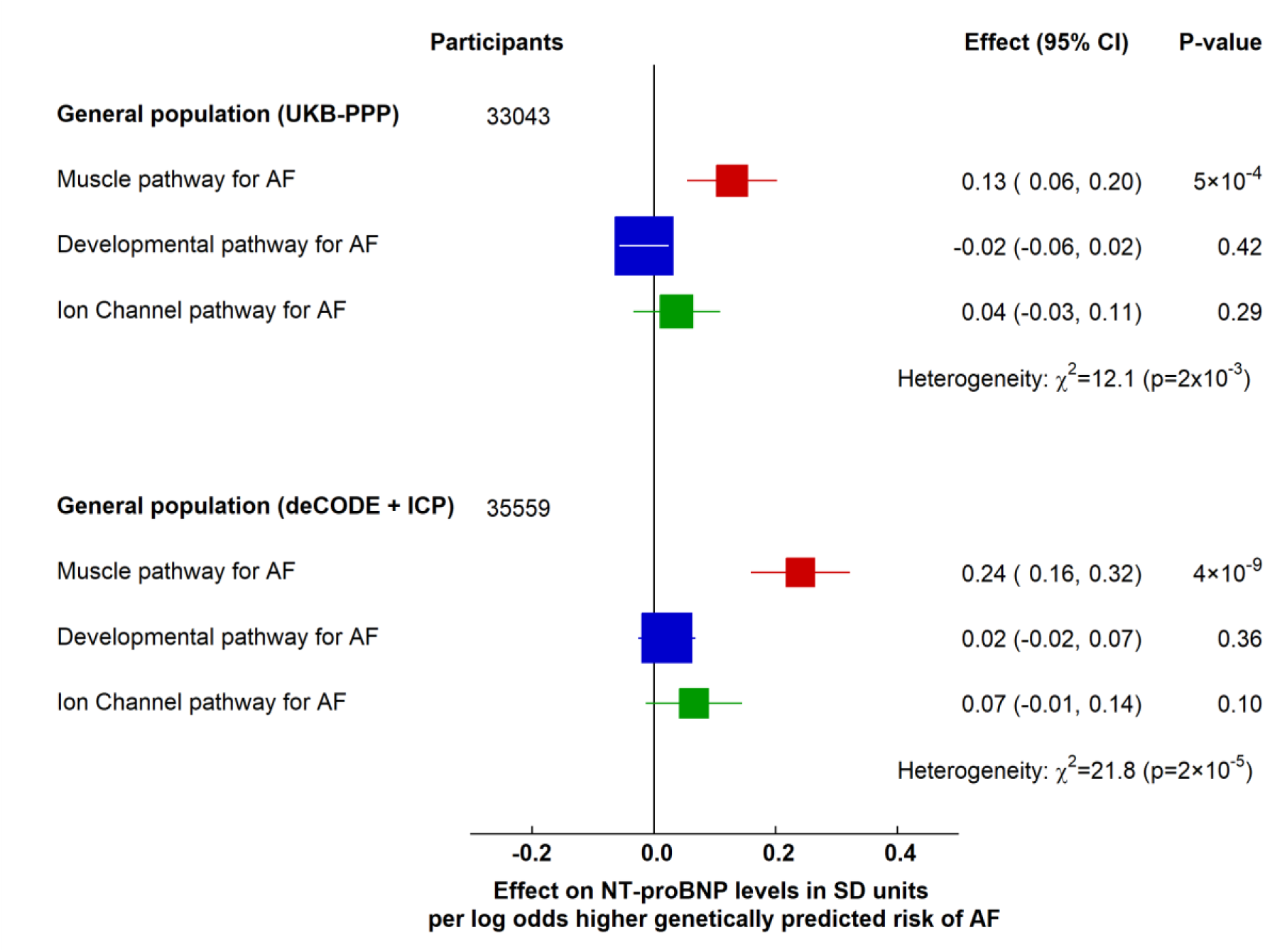
Effects of genetically predicted AF biological pathways on natriuretic peptide levels. Effect of AF pathways on N-terminal pro B-type natriuretic peptide (NT-proBNP) in standard deviation (SD) units in 33,043 British individuals from the UK Biobank Pharma Proteomics Project (UKB-PPP) and, in 35,559 Icelandic individuals from the deCODE and Icelandic cancer project (ICP). Boxes represent point estimates of effect per log odds higher genetically predicted risk of AF, with their size inversely proportional to variance and solid lines representing 95% confidence intervals (CI). Effect sizes were calculated using inverse variance weighted methods from summary GWAS data and heterogeneity using Cochran Q.

To assess the sensitivity of these findings to population characteristics, we examined the associations in a population of 3,301 healthy blood donors from the INTERVAL study^35^ (mean age 43 years) and in 3,394 older adults with risk factors for cardiovascular disease from the IMPROVE study^37^ (mean age 65 years). Consistent with UKB-PPP and deCODE + ICP, only the muscle pathway was associated with a higher level of NT-proBNP, with significant heterogeneity between pathways seen in both cohorts (*P =* 4 × 10^-3^ and *P* = 0.04 respectively; **Supplementary** Figure 6). The 142-variant set for overall risk of AF was also associated with higher levels of NT-proBNP, but only in the UKB-PPP and deCODE + ICP cohorts (**Supplementary** Figure 7).

#### High sensitivity Troponins

To evaluate whether changes in LA function and biomarkers of LV dysfunction were linked to myocardial injury, we investigated associations with hsTnI (multi-ancestry population of 48,115 individuals^39^ and in a separate multi-ancestry population of 14,336 individuals free from prevalent coronary heart disease or heart failure^40^) and hsTnT (24,617 multi-ancestry individuals free from prevalent coronary heart disease or heart failure^40^) using GWAS summary data.

We found no differences between the biological pathways in terms of their associations with hsTnI or hsTnT (*P* for heterogeneity 0.14 and 0.36 for the hsTnI cohorts, and 0.85 for the hsTnT cohort; **Supplementary Figure 8**).

The 142-variant set was not associated with higher hsTnI levels in the general population but was associated with a slightly higher level of both TnI and TnT in multi-ethnic populations free from prevalent coronary heart disease or heart failure (**Supplementary Figure 9**).

Overall, these data provide further evidence that different mechanisms underlying the genetic risk of AF are associated with distinct cardiac phenotypes and suggest that the muscle pathway is associated with biomarkers of LV dysfunction, but not myocardial injury, in addition to left atrial dysfunction.

### Genetic predisposition to AF conferred by mechanistic pathways is associated with different risks of cardioembolic, but not small vessel ischaemic stroke

We next investigated whether different pathways of genetically-predicted risk for AF may be associated with a different risk of cardioembolic stroke, for which AF is a significant risk factor, but not with small-vessel stroke (as a negative control) due to its differing risk factor profile. We used summary statistics from the GIGASTROKE meta-analysis employing the TOAST classification system^42^ to categorise acute stroke subtypes.

In analyses limited to individuals of European ancestry (comparable to the population in which the AF GWAS was undertaken^12^), the three pathways were differentially associated with risk of cardioembolic stroke (*P* for heterogeneity 0.04; **Figure 4**). Specifically, the ion channel pathway was associated with a lower risk of cardioembolic stroke, most profoundly when compared with the developmental pathway (*P* = 0.01). In the larger multi-ancestry GIGASTROKE meta-analysis, we were similarly able to replicate the differences in the risk of cardioembolic stroke across the different pathways (*P* for heterogeneity 7 ×10^-3^), again with a lower risk conferred by the ion channel pathway (OR for cardioembolic stroke, 1.66; 95% CI 1.43, 1.93 per log odds higher genetically predicted risk of AF), particularly when compared with the developmental pathway (*P* = 1 ×10^-3^; **Figure 4**). By contrast, none of the individual pathways was associated with small-vessel ischaemic stroke, with no significant heterogeneity among the estimates (*P* for heterogeneity 0.20; **Supplementary Figure 10**). Consistent with previous MR studies of AF and stroke^46, 47^, the 142-variant set for overall AF susceptibility was associated with a higher risk of cardioembolic stroke, but not with small-vessel ischaemic stroke (**Supplementary Figure 11**).

**Figure 4.**
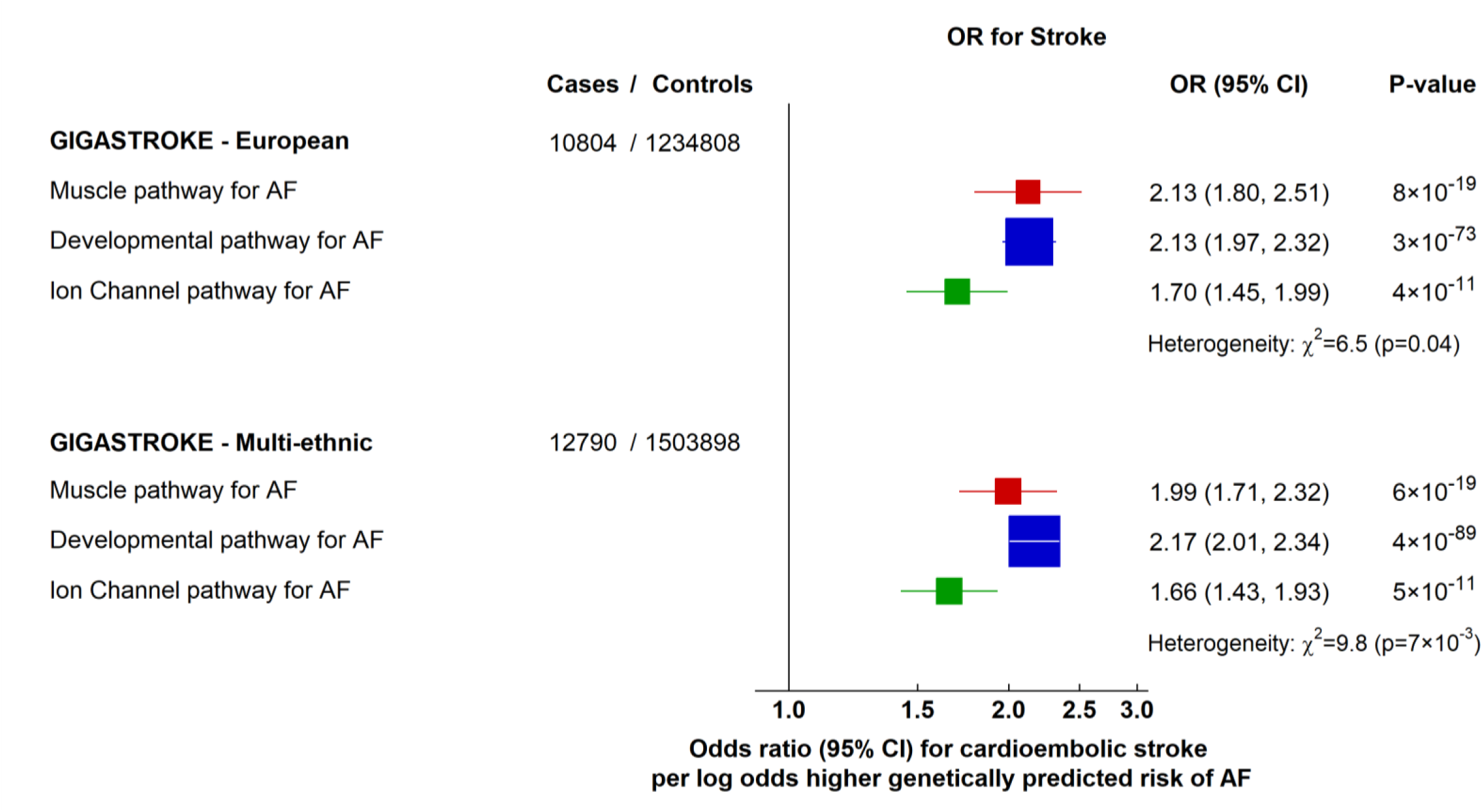
Effects of genetically predicted AF biological pathways on cardioembolic stroke. Odds ratio (OR) for developing types of ischaemic stroke in the GIGASTROKE dataset. Boxes represent point estimates of effect per log odds higher genetically predicted risk of AF, with their size inversely proportional to variance and solid lines representing 95% confidence intervals (CI). ORs were calculated using inverse variance weighted methods from summary GWAS data, and heterogeneity using Cochran Q.

Overall, these findings suggest that different mechanisms underlying the genetic risk of AF are not only associated with distinct intermediate phenotypes but also with different risks of cardioembolic stroke.

### Sensitivity Analyses

#### Investigations for pleiotropic effects within the pathways

Sensitivity analyses conducted for each phenotype did not suggest pleiotropy, and indicated effect estimates were robust to methodological assumptions. Further details are available in the **Supplementary Results** section and **Supplementary Table 10**. Funnel plots to visualise the effects of each variant on risk of cardioembolic stroke within each AF score are shown in **Supplementary Figures 12 - 14** and ‘leave-one-out’ analyses suggested results were robust to the exclusion of individual genetic variants (**Supplementary Figures 15 - 17**).

#### Potential effects of pathway-associated intermediate phenotypes on risk of cardioembolic stroke

We performed analyses for cardioembolic stroke adjusted for the effects of the variants on the intermediate phenotypes, grouped by ECG parameters, markers of LA size and function, and then NT-proBNP and LV ejection fraction. Estimates were comparable before and after adjustment (**Supplementary Tables 11 - 13**), suggesting that associations between the variants and intermediate phenotypes examined in this study are unlikely to fully explain the differences in association with cardioembolic stroke.

## DISCUSSION

Our findings indicate that distinct biological pathways underlying the genetically-predicted risk of AF are differentially associated with atrial and ventricular phenotypes, and, importantly, with the risk of cardioembolic stroke. This study not only validates the heterogeneous nature of the disease processes predisposing to AF, but highlights the potential for improved risk stratification and more targeted therapeutic strategies.

### Biological pathways for AF

A variety of biological pathways underpinning the development of AF have been identified using clinical, laboratory and animal methods^1, 2^. Commensurate with early understanding of AF as a primarily ‘electrical’ disease, initial linkage analyses identified ion-channel genes as potentially relevant in the pathogenesis of AF^3, 48^. More recently, large GWAS have identified many common genetic variants underlying AF susceptibility and have used an array of *in-silico* approaches to suggest their relevance to distinct biological pathways^12^. Within this framework however, combining information across the genome without biological considerations could obscure important mechanistic distinctions, prompting us to investigate the relevance of discrete biologically-informed genetic pathways associated with AF risk.

Our findings suggest that different AF-related mechanisms have different effects on biomarkers, intermediate phenotypes, and risk of cardioembolic stroke, which may partially explain the heterogeneity seen in observational studies linking ECG ^16–18^, cardiac imaging parameters^19–22^, and natriuretic peptides^49, 50^ with risk of AF.

### Atrial and ventricular substrates

Changes in atrial and ventricular substrates in the context of AF are complex and have been well described^1, 51^, with a number of intermediate phenotypes affecting both atria and ventricles being relevant to AF (e.g. P-wave duration or QT interval^16, 17^, LA volume^52^ or emptying fraction^53^, and LV dysfunction^22^). We initially investigated whether genetic variation associated with risk of AF and representing three distinct pathways (ion-channel, muscle structure and function, and developmental pathways) had different effects on intermediate phenotypes in individuals in sinus rhythm.

Out of the surface ECG parameters, most strikingly, the muscle and the ion-channel pathways had directionally opposite associations with the PR interval. Strong associations between the pathways with P-wave duration were not seen in our study and thus the heterogeneity seen for associations with the PR interval persisted even after adjusting for P-wave duration. This may suggest that variations in AV node transit time rather than measures of atrial depolarisation reflect different biological pathways to AF and related outcomes. However, ECG parameters in UK Biobank are based on automated ECG recordings and measuring P-waves using automated ECG algorithms has considerable challenges given their low amplitude and variability across different ECG leads^54, 55^, so it is possible that the PR interval associations simply more accurately reflect atrial electrical parameters or include atrial repolarisation which the P-wave duration may not.

Observational data have suggested that both short and long P-wave durations and PR intervals are associated with a higher risk of AF^17, 18^, and Mendelian randomisation studies have suggested shorter atrial ECG parameters are associated with a higher risk of AF, commensurate with the effects of the ion-channel pathway^56^. Our findings are consistent with data linking an atrial cardiomyopathic substrate to slower atrial conduction^57–59^, whereas shorter PR intervals may reflect shortening of atrial action potential duration (as seen in those with AF^60^) and more rapid conduction. Further investigation would be needed to establish whether these insights also provide information on the mechanisms underpinning AF development (e.g. re-entrant versus multiple wavelets, or trigger-dependent versus substrate dependent).

Our study showed differing associations of the AF-associated biological pathways with NT-proBNP, with the muscle pathway being associated with higher plasma levels across all cohorts, including in seemingly healthy, younger subjects. We also observed associations between the muscle and ion-channel pathways and QRS interval duration on ECG, and that the muscle pathway was associated with a lower indexed LV stroke volume. The muscle pathway also had the strongest associations with lower left atrial function, particularly in the passive emptying phase, which may indicate associations with LV diastolic dysfunction more generally.

Associations between NT-proBNP and AF have been reported widely in the literature alongside a consistently increased risk of stroke^24, 25^. Our previous work also indicated that people with “lone” AF continue to show evidence of subtle ventricular dysfunction and impaired myocardial energetics on cardiac magnetic resonance imaging/spectroscopy, despite normalisation of rhythm ∼6 months post-ablation, consistent with the presence of a subclinical cardiomyopathy underpinning AF^22^.

Our analyses did not support any consistent difference in the effect of AF-associated pathways on hsTnI/T levels in a general population, or separately in a cohort of individuals free from coronary artery disease or heart failure. In subjects with AF, hsTnI/T levels have been shown to be relevant to risk-stratification for future cardiovascular outcomes^25–27^. Whilst the muscle pathway in particular was associated with markers of LA and LV dysfunction, this does not necessarily reflect the presence of myocardial injury and cell death. Other findings suggest that this relationship may be accounted for, at least in part, by concomitant coronary disease^61^.

### Different risks of stroke based on different pathways to AF

Our study suggests that genetic variants representing different pathways associated with risk of AF can confer differing risks of cardioembolic stroke. There is evidence suggesting that structural or electrical changes within the atria may lead to thromboembolism and AF independently^52, 62–64^.

Furthermore, missense mutations in genes associated with changes in atrial structure confer an increased risk of both AF and stroke^8, 9, 65^. This has given rise to the hypothesis that AF and stroke may be the distinct endpoints of a cardiomyopathic substrate^15, 66–69^. In our study, the same risk of AF underpinned by different biological pathways was associated with differences in the risk of cardioembolic stroke.

Whilst we demonstrated associations between the ion channel pathway and ECG parameters, as well as the muscle pathway and some of the known intermediate phenotypes for atrial and ventricular cardiomyopathy, the developmental pathway for AF was associated with a higher risk of stroke in the absence of an ‘obvious’ atrial phenotype as assessed by conventional biomarkers. Although the function of the specific variants included in the developmental pathway (*PITX2*, *NKX2-5*, *GATA 4* and *TBX5*) is only partially known^4^, these genes may impact cardiac development in early life, with evidence that missense mutations in *TBX5* can lead to profound atrial transcriptional alteration and AF^70^. Investigation of more sophisticated biomarkers may yield more insights into understanding how this pathway impacts AF and stroke risk.

Our study provides insights into disease aetiology that may allow better risk profiling of patients and help inform treatment pathways. Further studies are needed to understand if individuals with AF who have risk profile patterns associated with different biological mechanisms may derive more benefit from specific interventions.

### Limitations

To define our biological pathways, we used the current largest GWAS for AF and its multi-modal *in-silico* gene and pathway annotations, and selected variants meeting GWAS significance^12^. Pathway annotation is complex and based on existing knowledge of functions of genes and thus limited by new scientific discovery. Whilst the associations observed with different intermediate phenotypes validates the different pathways represented to some extent, we did not comprehensively test distinct biological pathways as a whole, but rather specific variants within these pathways known to be associated with AF.

The GWAS for AF used for genetic variant selection were conducted in participants with European ancestry, and outcomes were also chiefly examined in studies of European ancestry, so further studies would be needed to assess the generalisability of our results across more diverse ethnicities.

### Conclusions

Genetic variants representing distinct biological pathways for AF susceptibility have differing effects on intermediate cardiac electrical, structural and functional phenotypes, extending to the ventricle, and risk of cardioembolic stroke. These findings provide a better understanding of the aetiological heterogeneity driving the development of AF and may offer a route to genetically-informed treatment strategies.

## ACKNOWLEDGEMENTS AND SOURCES OF FUNDING

This work was supported by the British Heart Foundation (grant numbers FS/17/17/32438 [PRG, BC], FS/20/15/34920 [CFC, JCH], RG/16/12/3245 and CH/12/3/29609 [BC], and FS/14/55/30806 and CH/1996001/9454 [JCH]). The authors also acknowledge support from: the European Union (grant agreement 633196 - CATCH ME); BHF Centre for Research Excellence, Oxford, National Institute of Health Research (NIHR) Oxford Biomedical Research Centre, and Nuffield Department of Population Health, University of Oxford, UK. British Heart Foundation.

The funders had no role in study design, data collection or analysis, preparation of the manuscript, or decision to publish.

## DATA AVAILABILITY STATEMENT

Data are available from UK Biobank (http://biobank.ndph.ox.ac.uk/showcase/) in accordance with their published data access procedures. All other data are publicly available or were obtained from corresponding authors of studies as detailed in the **Methods** section and **Supplementary Table 5**.

